# Predicting Walking Capacity Outcomes After Moderate to High Intensity Locomotor Training in Chronic Stroke

**DOI:** 10.1101/2024.08.24.24312537

**Authors:** Christina Garrity, Darcy S. Reisman, Sandra A. Billinger, Katie A. Butera, Pierce Boyne

## Abstract

**Purpose:** Moderate-to-high intensity locomotor training (M-HIT) is strongly recommended in stroke rehabilitation but outcomes are variable. This study aimed to identify baseline clinical characteristics that predict change in walking capacity following M-HIT in chronic stroke.

**Methods:** This analysis used data from the HIT-Stroke Trial (N=55), which involved up to 36 sessions of either moderate or high intensity locomotor training. A prespecified model assessed how well baseline motor impairment (Fugl-Meyer lower limb motor scale [FM-LL]), comfortable gait speed (CGS), and balance confidence (Activities Balance Confidence scale [ABC]), independently explain changes in 6-minute walk distance (Δ6MWD), while controlling for treatment group. Exploratory analysis tested additional baseline covariates using the all-possible regressions procedure. The prognostic value of each potential covariate was assessed by its average contribution to the explained variance in Δ6MWD (Δ pseudo-R^2^).

**Results:** With the prespecified model, 8-week Δ6MWD was significantly associated with baseline FM-LL (β=5.0 [95% CI: 1.4, 8.6]) and ABC (β=0.7 [0.0, 1.4]), but not CGS (β=-44.6 [-104.7, 15.6]). The exploratory analysis revealed the top 7 covariates with the highest mean Δ pseudo-R^2^ were: FM-LL, pain-limited walking duration, ABC, the use of an assistive device, fatigue, depression, and recent walking exercise history >2 days per week.

**Conclusions:** On average, participants with less motor impairment and higher balance confidence have greater walking capacity improvements after M-HIT in chronic stroke. Additional negative prognostic factors may include pain-limited walking duration, use of an assistive device, fatigue, depression, and recent walking exercise but these exploratory findings need to be confirmed in future studies.

## INTRODUCTION

Current clinical practice guidelines for improving walking capacity (a clinic-based measure of walking endurance to combine speed with longer distance walking) in chronic stroke recommend using moderate-to-high intensity locomotor training (M-HIT). This approach focuses on walking training at faster than comfortable gait speed (CGS) and with a goal of reaching aerobic training intensities greater than 39% heart rate reserve.^1,2^ M-HIT has been shown to be effective in improving walking capacity measures, such as six minute walk distance (6MWD), in chronic stroke.^3^ However, there is variability in patient outcomes, beyond that expected from random day-to-day variability and measurement error.^3-5^

This variability makes it difficult to know how much an individual may benefit from M-HIT. Accurately predicting walking outcomes at the outset of an episode of care could help with setting goals, monitoring progress, and individualizing rehabilitation care plans.^6^ Many studies have attempted to identify baseline characteristics that may estimate future potential for change in walking outcomes. However, there is large heterogeneity between studies in which baseline characteristics are tested, how they are modeled, and how they are reported. The baseline characteristic most consistently shown to predict walking capacity outcomes in chronic stroke has been baseline CGS.^7-12^ Other variables that may also contribute but have not been as well studied include balance,^9,13,14^ lower extremity motor function,^9,15-17^ time since stroke,^5,18^ steps per day,^13^ lesion location,^5^ and age.^9,18^

Some of these potential prognostic variables are correlated with CGS, making it possible that some variance in outcomes explained by CGS could be attributable to one or more of these other covariates. No previous studies have aimed to assess how well baseline CGS independently contributes to changes in walking capacity when controlling for these other correlated baseline clinical characteristics. In addition, prognostic research in general is typically done on an exploratory basis, with high potential for false positive findings.^19^ For example, prognostic variables are often identified by testing a multitude of potential predictors, which increases the risk of overfitting the model to the random error specific to an individual dataset. Thus, the recommended approach for predicting clinical outcomes is to test a prespecified multivariable model when possible.^19^

A recent multi-center trial comparing moderate versus high intensity gait training in chronic stroke (HIT-Stroke Trial)^20^ provides a new opportunity to assess baseline predictors of walking capacity outcomes following moderate or high intensity locomotor training.^21^ Here, we report the results of a prespecified HIT-Stroke Trial analysis that aimed to assess the prognostic value of baseline CGS, lower extremity Fugl-Meyer (FM-LL) motor scores, and balance confidence for predicting walking capacity changes (Δ6MWD; see ClinicalTrials.gov record NCT03760016 for this prespecified analysis plan). These measures were selected because they all represent different mobility constructs that have shown potential for predicting response to gait rehabilitation in separate studies (CGS,^7-11,15,18,21^ motor function,^15-17^ and balance^9,13,14^). Importantly, no previous studies have tested all three constructs in the same multivariable prognostic model to better understand how each construct may be independently explaining variance in walking outcomes. Since the HIT-Stroke Trial did not obtain any measures of balance, we substituted the available measure of balance confidence. To further inform future studies, we also explored the prognostic value of other clinically available baseline characteristics.

## METHODS

This analysis used all available data from the 55 participants in the HIT-Stroke Trial. All study participants underwent 45-minute training sessions that included both overground and treadmill walking exercise, 3 times a week for 12 weeks. Participants were randomized into either the moderate intensity aerobic training (MAT) group or the high intensity interval training (HIIT) group. The MAT group (N=28) performed continuous walking with speed adjusted to maintain an initial target heart rate of 40% heart rate reserve (HRR), which was incrementally increased 5% every 2 weeks up to 60% HHR as tolerated. The HIIT group (N=27) performed repeated 30 second bursts at maximal safe walking speeds alternated with 30-60 second passive rest (standing or seated), targeting above 60% HRR. The primary 6MWD outcome was assessed by blinded raters at baseline (PRE) and after 4, 8, and 12 weeks of training (POST-4WK, POST-8WK and POST-12WK). The current HIT-Stroke Trial analysis tested the prognostic value of different baseline participant characteristics for predicting change in walking capacity (Δ6MWD) following HIIT or MAT (i.e. moderate to high intensity locomotor training), while controlling for the effects of treatment intensity group.

### Setting and Participants

Participants were recruited from the community and provided written informed consent prior to participation. Inclusion criteria were: (1) age of 40 to 80 years at the time of consent, (2) single stroke for which the participant sought treatment 6 months to 5 years prior to consent, (3) walking speed of 1.0 m/s or less, (4) ability to walk 10 m over ground with assistive devices as needed and no continuous physical assistance from another person, (5) ability to walk at least 3 minutes on a treadmill at 0.13 m/s (0.3 mph) or faster, (6) stable cardiovascular condition (American Heart Association class B), and (7) ability to communicate with investigators, follow a 2-step command, and correctly answer consent comprehension questions.

Exclusion criteria were: (1) exercise testing uninterpretable for ischemia or arrhythmia, (2) evidence of significant arrhythmia or myocardial ischemia on a treadmill ECG-graded exercise test in the absence of a more definitive negative result from clinical testing, (3) hospitalization for cardiac or pulmonary disease within the past 3 months, (4) implanted pacemaker or defibrillator; (5) significant ataxia or neglect (National Institutes of Health Stroke Scale item scores >1), (6) severe lower limb spasticity (Ashworth Scale scores >2 of 4 for knee flexion, knee extension, or ankle dorsiflexion), (7) recent history of illicit drug or alcohol misuse or significant mental illness, (8) major poststroke depression (Patient Health Questionnaire score ≥10) in the absence of depression management by a health care professional, (9) currently participating in physical therapy or another interventional study, (10) recent botulinum toxin injection to the paretic lower limb (<3 months previously) or planning to have it in the next 4 months, (11) foot drop or lower limb joint instability in the absence of an adequate stabilizing device (eg, ankle foot orthosis), (12) clinically significant neurologic disorder other than stroke, (13) unable to walk outside the home prior to stroke, (14) other significant medical condition likely to limit improvement or jeopardize safety, (15) pregnancy, and (16) previous exposure to fast treadmill walking in the past year.

### Data Collection

The primary walking capacity outcome was 6-minute walk distance (6MWD).^22^ Baseline measures included in the analysis as potential predictors of Δ6MWD were those that could be performed in a traditional clinical rehabilitation setting. To address expected collinearity among related baseline characteristics in the exploratory analysis, these measures were first grouped by construct. The constructs and their representative covariates were:

#### (1) Motor function

FM-LL total motor score^23^ and FM-LL synergy score, which included only volitional movement scores (not reflexes or coordination);

#### (2) Gait speed

CGS (10-meter walk test),^24^ fastest gait speed, CGS dichotomized by ≥ 0.4 m/s^7^, CGS dichotomized by ≥ 0.5 m/s^25,26^, gait speed reserve (fastest gait speed minus CGS);

#### (3) Walking independence

functional ambulation category (FAC) > 3 (can ambulate independently on level surfaces), FAC > 2 (can ambulate on level surfaces with or without supervision);^27^

#### (4) Assistive device use

use of any handheld assistive device, use of a weightbearing assistive device (i.e. more than a single point cane);

#### (5) Pain

those who reported pain with walking were asked “Does pain limit how far or how long you can walk?” (yes/no), “How severe is the pain typically while you are walking?” (0-10), “Does the pain increase the more you walk?” (yes/no);

#### (6) Recent exercise history

“In the past month, how often have you done aerobic exercise?” (< 1 time per week, 1-2 times per week, > 2 times per week), how was exercise performed (seated and/or walking), “Was exercise vigorous enough to make you sweat?” (yes/no).

The remaining baseline measures considered as potential covariates only included a single variable per construct. These included the Activities-specific Balance Confidence Scale (ABC) as a measure of balance confidence,^28,29^ the EuroQOL-5D-5L misery score (EQ-5D) as a measure of quality of life (higher scores indicate lower quality of life),^30^ the Patient Reported Outcomes Measurement Information System (PROMIS) Fatigue Scale as a measure of fatigue,^31,32^ and the Patient Health Questionnaire-9 (PHQ-9) as a measure of depression.^33^ All the above measures with references have been shown to be reliable and valid in persons with stroke or chronic health conditions.

### Statistical Analysis

All prognostic analyses were done with an intent-to-treat approach using a linear model for 6MWD. All models included fixed effects for group (HIIT or MAT), categorical testing time point (PRE, POST-4WK, POST-8WK, POST-12WK) and group-by-time interaction, with unconstrained covariance between repeated measures within participants. Different analyses added to this model different baseline covariates and their interaction with time. To obtain a standardized measure of prognostic model accuracy, a pseudo-R^2^ statistic was obtained for each model and change time point. Pseudo-R^2^ was calculated as the squared correlation between observed and model-estimated Δ6MWD at a given POST time point.

### Primary prognostic analysis

The prespecified primary prognostic model included the following baseline covariates and their interactions with time: CGS, FM-LL motor score, and the ABC score. As in our previous analysis,^34^ POST-8WK was designated as the primary time point of interest since it allowed more intervention duration than POST-4WK, with less missing data than POST-12WK. To help interpret the magnitude of the estimated effects of this analysis, the coefficients were also scaled to a minimal clinically important difference (MCID) for the baseline covariate. This was done by multiplying the coefficients and their confidence intervals by the MCID. The selected MCID values were: FM-LL motor score^39^, 6 points;^35^ CGS, 0.10 m/s;^36^ ABC score, 15%;^37^ 6MWD, 20 meters.^38,39^

The overall modelling strategy described above assumed that the prognostic effects of a given baseline covariate were the same for both training intensity groups (HIT and MAT). This assumption was tested in a sensitivity analysis by adding covariate-by-group-by-time interactions to the primary model to estimate the covariate-by-time interactions (i.e. prognostic effects) within each group for comparison.

### Exploratory prognostic analysis

An exploratory analysis was performed to assess whether any important prognostic covariates may have been missed in the pre-specified prognostic model, by also considering the additional baseline clinical measures described in the ‘data collection’ section above. This exploratory analysis tested all possible combinations of the potential baseline covariates (i.e. the ‘all-possible regressions’ procedure^40^), with up to 5 covariates and their interactions with time in a given model. To avoid collinearity issues from including highly correlated baseline covariates in the same model, we selected a single covariate from each construct for this analysis. To partially contain the risk of overfitting, the covariates used for each construct were pre-selected based on previous studies as much as possible (see supplemental appendix for details). Finally, to derive a single ‘best model’ to maximally explain variance in Δ6MWD (for future validation), we identified the model with the highest (adjusted) Δ pseudo-R^2^ from the ‘all possible regressions’ procedure. A more detailed description of this exploratory analysis can be found in the supplemental appendix.

## RESULTS

Baseline participant characteristics are shown in Table 1. Out of the 55 participants that had baseline testing and were randomized, 53 had 6MWD data at POST-4WK, 47 at POST-8WK, and 42 at POST-12WK. Ten participants discontinued participation before POST-12WK due to events likely unrelated to the study (including but not limited to COVID-19 shutdown) and 3 participants discontinued due to treatment-related adverse events.^20^

**Table 1.** Baseline participant characteristics (N=55)

| **Table 1. Baseline participant characteristics (N=55)** |  |
| --- | --- |
| Age (years) | 62.4 ± 9.7 |
| Sex |  |
| Female | 19 (34.5) |
| Male | 36 (65.5) |
| BMI | 28.8 ± 4.9 |
| Years since stroke | 2.5 ± 1.3 |
| Hemorrhagic stroke | 21 (38.2) |
| Number of falls in last 3 months | 0.4 ± 0.8 |
| Lives alone | 9 (16.4) |
| Uses an orthotic | 21.0 ± 38.2 |
| ABC Score (0-100%) | 66.4 ± 19.5 |
| EuroQOL-5D-5L misery score (5-25) | 10.1 ± 2.8 |
| PROMIS-Fatigue scale (T-score) | 50.6 ± 7.6 |
| PHQ-9 (0-27) | 2.7 ± 3.2 |
| **Motor Impairment Construct** |  |
| Fugl-Meyer lower limb motor score | 23.4 ± 5.0 |
| Fugl-Meyer synergy score | 15.4 ± 4.2 |
| **Gait Speed Construct** |  |
| Baseline comfortable gait speed (m/s) | 0.64 ± 0.30 |
| Baseline fast gait speed (m/s) | 0.86 ± 0.45 |
| Comfortable gait speed ≥ 0.4 m/s | 41 (74.5) |
| Comfortable gait speed ≥ 0.5 m/s | 36 (65.5) |
| Gait speed reserve (m/s) | 0.22 ± 0.19 |
| **Assistive Device Construct** |  |
| Use of any assistive device | 34 (61.8) |
| Weightbearing assistive device | 6 (10.9) |
| **Walking Independence Construct** |  |
| Functional Ambulation Category >2 | 48 (87.3) |
| Functional Ambulation Category >3 | 26 (47.3) |
| **Pain Construct (n=54)\*** |  |
| Reported pain-limited walking duration | 8 (14.5) |
| Reported pain increases with walking | 6 (10.9) |
| Pain severity with walking (0-10) | 1.1 ± 2.0 |
| **Exercise History Construct (n=54)\*** |  |
| Exercise frequency >0 d/wk | 29 (52.7) |
| Seated exercise >0 d/wk | 7 (12.7) |
| Walking exercise >0 d/wk | 24 (43.6) |
| Exercise >0 d/wk and sweat producing | 16 (29.1) |
| Seated exercise >0 d/wk and sweat producing | 4 (7.3) |
| Walking exercise >0 d/wk and sweat producing | 14 (25.5) |
| Exercise frequency >2 d/wk | 16 (29.1) |
| Seated exercise >2 d/wk | 5 (9.1) |
| Walking exercise >2 d/wk | 11 (20.0) |
| Exercise >2 d/wk and sweat producing | 8 (14.5) |
| Seated exercise >2 d/wk and sweat producing | 3 (5.5) |
| Walking exercise >2 d/wk and sweat producing | 6 (10.9) |
| Values reported as mean ± SD or N (%). *One participant had missing values for the pain questions at baseline and a different participant had missing values for the exercise history questions. Abbreviations: SD, standard deviation; BMI, body mass index; ABC, Activities-specific Balance Confidence scale; PHQ, Patient Health Questionnaire; m/s, meters per second; d/wk, days per week |  |

### Primary prognostic analysis

The prespecified multivariable model (Figure 1, Table 2) found that greater Δ6MWD from PRE to POST-8WK was associated with higher baseline values for the FM-LL (β =4.99 [95% CI: 1.42, 8.56]) and ABC scale (β =0.72 [0.02, 1.42]), but not CGS (β =-44.55 [-104.68, 15.58]). The model explained 32.9% of the variance in Δ6MWD. Mean (SD) absolute error was 32 (21) meters for the POST-8WK time point. Results for other change time points are presented in Table 2. In the sensitivity analysis, when the primary model covariate associations were temporarily allowed to vary by treatment group (i.e. group-by-covariate-by-time interactions), the estimated 8-week Δ6MWD between-group differences were well below the 20-meter MCID and not statistically significant (see supplemental appendix). Therefore, no further analyses attempted to estimate prognostic effects separately by group.

**Table 2.** Prespecified multivariable model for predicting change in 6-minute walk distance (△6MWD) after moderate-to-high intensity locomotor training.

|  | POST-4WK | **POST-8WK\*** | POST-12WK |
| --- | --- | --- | --- |
| Model coefficients (95% CI) for baseline covariate-by-time interactions<br>($\Delta 6\text{MWD}$ associated with every one unit higher value of covariate) | | | |
| Fugl-Meyer lower limb motor score (0-34) | 3.4 (0.4, 6.3) | **5.0 (1.4, 8.6)** | 4.3 (-0.3, 8.8) |
| Comfortable gait speed (m/s) | -9.2 (-58.5, 40.2) | **-44.6 (-104.7, 15.6)** | -35.4 (-113.0, 42.2) |
| ABC score (0-100%) | 0.6 (-0.0, 1.1) | **0.7 (0.0, 1.4)** | 0.8 (-0.1, 1.7) |
| Coefficients (95% CI) from above rescaled to an MCID in the covariate<br>($\Delta 6\text{MWD}$ associated with a one MCID higher value of covariate) | | | |
| Fugl-Meyer lower limb motor score (MCID: 6 points) | 20.1 (2.1, 38.0) | **29.9 (8.6, 51.3)** | 25.5 (-1.7, 52.7) |
| Comfortable gait speed (MCID: 0.1 m/s) | -0.9 (-5.7, 4.0) | **-4.5 (-10.5, 1.6)** | -3.5 (-11.3, 4.2) |
| ABC score (MCID: 15%) | 8.5 (-0.0, 17.1) | **10.8 (0.3, 21.3)** | 11.5 (-2.0, 25.1) |
| Model accuracy |  |  |  |
| Pseudo-$R^2$<br>(squared correlation between predicted and observed $\Delta 6\text{MWD}$) | 35.9% | **32.9%** | 23.2% |
| Mean (SD) absolute error<br>($\Delta 6\text{MWD}$ meters) | 24 (22) | **32 (21)** | 39 (29) |
| *Primary time-point for analysis. Abbreviations: CI, confidence interval; WK, week; m/s, meters per second; MCID, minimal clinically important difference. |  |  |  |

**Figure 1.**
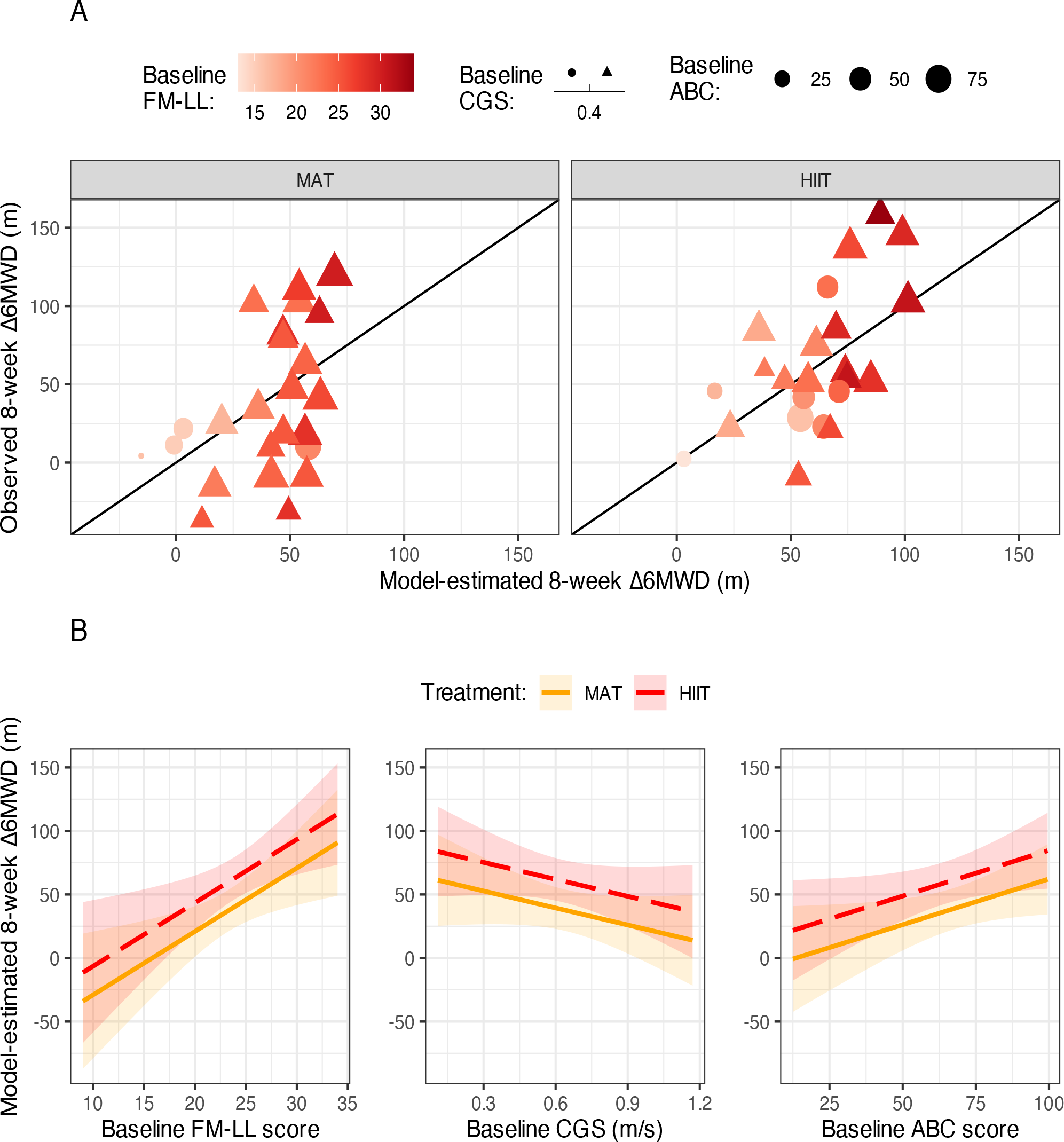
Results of primary model for predicting change in 6-minute walk distance (Δ6MWD). Panel A: Model predicted versus observed Δ6MWD. Lighter point color indicates greater baseline motor impairment. Point shape depicts baseline comfortable gait speed (CGS) ≤ 0.4 m/s (circles) or >0.4 m/s (triangles). Larger point size represents better baseline balance confidence. Panel B: Model predicted Δ6MWD for individuals with different values of a baseline covariate, when values for the remaining covariates in the model are at the mean of the study sample (FM-LL, 23.4 points; CGS, 0.64 m/s; ABC, 66.4%). Ribbon represents 95% confidence interval. Abbreviations: FM-LL, Fugl-Meyer lower limb motor score; ABC, Activities-specific Balance Confidence scale; m, meters; HIIT, high intensity interval training; MAT, moderate intensity aerobic training; m/s, meters per second.

All the estimated coefficients from the prediction model are provided in the supplemental Table S1. The prediction equation has also been integrated into a spreadsheet that can be used to obtain predicted 6MWD changes after 4, 8 and 12 weeks of either HIIT or MAT, by inputting baseline FM-LL, CGS and ABC scores (see final tab in supplemental tables file).

### Exploratory prognostic analysis

The exploratory analysis revealed the top 7 covariates with the highest mean Δ pseudo-R^2^ were: FM-LL, pain-limited walking duration, ABC, the use of an assistive device, PROMIS-Fatigue scale, PHQ-9, and recent walking exercise history >2 days per week (Figure 2). Each of these covariates had significantly (p<.05) higher Δ pseudo-R^2^ than all covariates below it, except the PROMIS-Fatigue scale was not significantly different from the PHQ-9 (see supplementary table S3). Positive T-statistics for FM-LL and ABC showed that higher values of these covariates were associated with greater Δ6MWD, while negative T-statistics for pain-limited walking duration, the use of an assistive device, PROMIS-Fatigue scale, PHQ-9, and recent walking exercise history indicated that these covariates were associated with lesser Δ6MWD (Figure 2).

**Figure 2.**
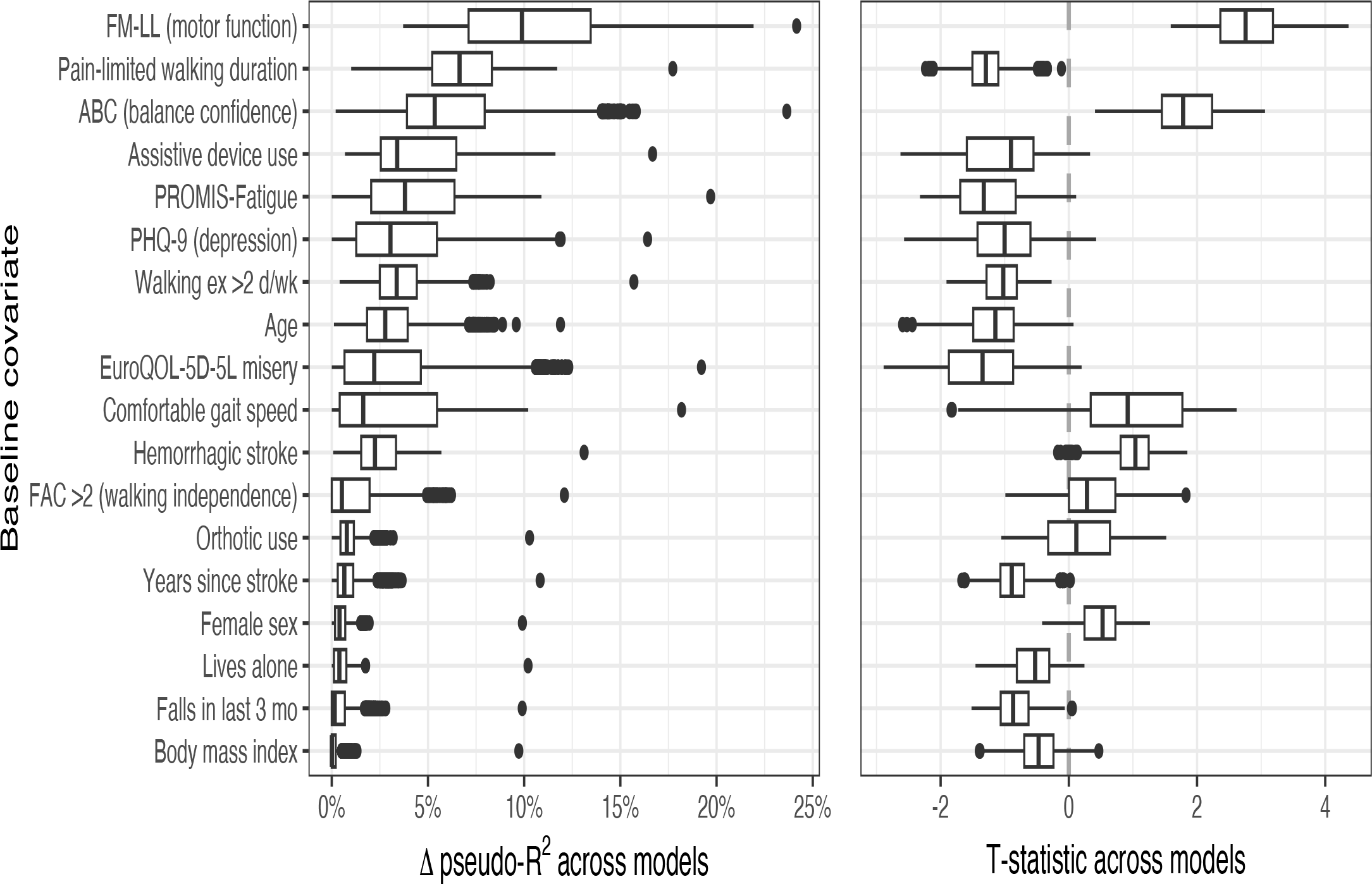
Prognostic value comparisons between baseline covariates. Left: Boxplots show the distribution of Δ pseudo-R^2^ across the 3,214 models involving each covariate, to depict which construct(s) explain the greatest variance in 6-minute walk distance change (Δ6MWD). Right: Boxplots show the distribution of T-statistics, primarily to depict whether each covariate was associated with greater (positive T-statistic) or lesser (negative T-statistic) Δ6MWD. Both: Values were averaged across POST-4WK, POST-8WK and POST-12WK change time points within each model. Abbreviations: FM-LL, Fugl-Meyer lower limb motor scores; ABC, Activities Balance Confidence Scale; PHQ, Patient Health Questionnaire; ex, exercise; d/wk, days per week; FAC, Functional Ambulation Category; mo, months; WK, week.

From the ‘all-possible regressions’ procedure, we also identified the single model with the highest adjusted pseudo-R^2^ using up to 5 baseline covariates: FM-LL + ABC + Age + Pain-limited walking duration + Walking exercise >2 d/wk. This model explained 46.0% of the variance in Δ6MWD on average across the POST time points. Coefficients from this model are provided in supplementary table S5.

## DISCUSSION

This study used a prespecified prediction model to better understand how baseline motor function, gait speed, and balance confidence each independently contribute to the variance in walking capacity outcomes following M-HIT in chronic stroke. Better motor function and better balance at baseline were each found to be independent predictors of greater walking capacity gains. This study also explored the prognostic value of additional covariates by assessing the average improvement in model accuracy attributable to each covariate across all possible multivariable models (with up to 5 baseline covariates). Results further confirmed the prognostic value of baseline motor function and balance confidence, while suggesting other potentially prognostic baseline covariates, including participant report of pain limiting walking duration, the use of an assistive device, fatigue, depression, and recent exercise history.

Motor function, as measured by FM-LL, was consistently shown to be the strongest prognostic covariate in both the pre-specified model and in the exploratory analysis. Thus, stroke survivors with greater baseline motor function can be expected to have greater walking capacity improvements after M-HIT, on average. This supports previous findings suggesting that baseline motor function may reflect an individual’s capability for gains in walking capacity.^15-17^

Previous studies have also shown that better baseline balance (e.g. Berg Balance Scale) may predict greater outcome improvements in response to M-HIT.^9,13,14^ As the HIT-Stroke Trial did not include a balance measure, we assessed the prognostic value of balance confidence (ABC scale), which has not been previously evaluated. In both the pre-specified model and in the exploratory analysis, greater baseline balance confidence significantly predicted greater walking capacity gains. Future studies are needed to determine whether balance confidence explains variance in Δ6MWD above and beyond that of balance.

Interestingly, this study found that baseline CGS does not independently contribute to M-HIT walking capacity outcomes when controlling for baseline motor function and balance confidence. On the surface, this may seem to conflict with previous chronic stroke studies, which have repeatedly found faster baseline CGS to be associated with better walking capacity gains.^7-11,18^ However, those prior associations were not controlled for other variables.^4,6-10^ When we mimicked those analyses by testing CGS as a lone predictor of walking capacity gains, we also similarly observed positive associations (e.g. β=36.7 for POST-8WK Δ6MWD; Table S6). Then, when controlling for motor function and balance confidence, the regression coefficients for CGS shrunk (e.g. from 36.7 to -44.6 for POST-8WK Δ6MWD; Table S6 and Table 2). This indicates that the positive association between CGS and walking capacity change observed in previous studies was likely confounded by motor function and/or balance confidence. Our exploratory analysis also indicated that motor function and balance confidence were significantly better predictors of walking capacity gains than CGS.

Yet CGS is still a highly recommended clinical measure in chronic stroke^1^ and is sometimes used in clinical research as a binary prognostic variable, dichotomized at 0.4 or 0.5 m/s (e.g. for stratified randomization).^7,9,10^ Therefore, it may still be helpful to know which baseline CGS variable(s) show the strongest relationships with walking capacity changes, even if those changes can be better explained by motor impairment and balance confidence. When multiple approaches for analyzing CGS were compared, CGS ≥ 0.4 m/s was a better predictor of walking capacity change than all other covariates in the CGS construct, including CGS as a continuous variable. This is atypical since dichotomous measures are less statistically powerful than continuous measures,^41^ which suggests that 0.4 m/s could be a biologically relevant CGS threshold. Interestingly, dichotomizing CGS at 0.4 m/s also explained significantly more variance in Δ6MWD than dichotomizing at 0.5 m/s.

Our exploratory variable importance analysis also provides additional insights into the relative importance of other baseline covariates for explaining variance in walking capacity gains after M-HIT. In addition to FM-LL and ABC, the 5 baseline covariates contributing the most to the explained variance in Δ6MWD were pain-limited walking duration, use of an assistive device, fatigue, depression, and recent walking exercise greater than 2 days per week. Each of these 5 covariates had a negative association with Δ6MWD, meaning that higher values predicted lower walking capacity gains following M-HIT. These findings for fatigue and depression are in alignment with previous literature.^42,43^ But, previous predictive models investigating walking capacity recovery following stroke have not tested the prognostic value of pain-limited walking duration, use of an assistive device, or recent walking exercise.

It seems particularly remarkable that the prognostic value of *pain* has not been previously tested in this context, since pain is a common experience post-stroke^44-46^ that has been associated with restricted mobility and lower self-reported quality of life, health status, and recovery.^46^ Robust evidence also indicates that pain has both sensory and motor consequences that impact movement.^47-50^ Thus, while a novel finding, it is not necessarily surprising that pain-limited walking duration explained significantly more variance in walking capacity change than any other baseline covariate except motor function in this study. This highlights the need for more routine and comprehensive pain measurement in stroke research. Future studies should consider assessing multiple pain characteristics (e.g., duration, location, severity, quality), as well as pain severity during movement and pain interference with daily activities. The current findings could also implicate pain as a potential treatment target to enhance outcomes, for instance, if the lower average walking capacity gain we observed among participants reporting pain-limited walking duration was *caused* by the pain. We hypothesize that more routine evaluation and management of pain among individuals following stroke could enhance responsiveness and/or participation in walking rehabilitation.

Using an assistive device and recent exercise engagement were also found to have prognostic value in explaining variance in walking recovery. We suspect that use of an assistive device may predict lower Δ6MWD because it may be related to lower balance confidence and/or pain, which this study already identified as being associated with lower Δ6MWD. Additionally, we speculate that recent walking exercise may predict lower Δ6MWD because individuals who were previously exercising at baseline may have already utilized some of their potential for exercise-mediated walking capacity gains.

In addition to evaluating the relative importance of different baseline covariates, our exploratory analysis also derived a single ‘best’ model to most accurately predict walking capacity gains after M-HIT. This model included FM-LL, ABC, age, pain-limited walking duration, and recent walking exercise greater than 2 days per week. Together, these covariates explained 46% of the variance in walking capacity gains, which is better than other previous models in chronic stroke that similarly used baseline clinical measures to predict walking outcomes (R^2^ = 25-37%).^5,51,52^ Thus, our derived model may be useful for guiding future studies. However, this model needs to be validated in another sample before it can be recommended for use, since its prognostic accuracy could have been falsely inflated by overfitting to the current data.^19^

### Strengths and Limitations

A strength of this analysis is that the primary prediction model was prespecified, which reduced the risk of overfitting.^19^ Another strength was that our multivariable models allowed for better evaluation of how each baseline covariate independently contributes to explained variance in walking outcomes. The exploratory analysis also provided a novel assessment of relative covariate importance and derived a preliminary model to optimize prediction accuracy while considering ‘all-possible regressions’.

This study was limited to the available baseline measures that were collected in the HIT-Stroke Trial, which did not include a performance-based balance measure. Thus, future prognostic studies should aim to include both perceived and performance-based balance measures to identify how different balance-related constructs may independently explain variance in walking outcomes. Similarly, pain measures included in this study provide only a general indication of a participant’s pain experience. Comprehensive evaluation of multiple pain characteristics (e.g., severity, quality, duration, interference, movement-evoked) is necessary to better understand the overall impact of pain on walking recovery following stroke. Another limitation of this study is that the results may only generalize to chronic stroke survivors who can ambulate at least 10 meters without physical assist and have other characteristics within the range of the study sample. For example, it is possible that stroke chronicity was a weaker predictor of walking outcomes in this study versus previous studies^5,18^ because we had a narrower eligibility range of 6 months to 5 years post-stroke. Lastly, these findings may or may not be specific to M-HIT, as we did not have another control group to test whether these covariates predict differential response to M-HIT versus other treatment approaches or no intervention.

## CONCLUSIONS

In a pre-specified analysis, motor function, CGS and balance confidence together explained 33% of the variance in walking capacity gains after 8 weeks of M-HIT. Greater motor function and balance confidence were each associated with significantly greater Δ6MWD gains, while CGS was not an independent predictor after accounting for these two variables. Our exploratory analysis found pain-limited walking duration, the use of an assistive device, fatigue, depression, and recent exercise history to also be prognostic, and the best single model was able to predict 46% of the variance in walking capacity gains. Future studies are needed to validate our exploratory results and further improve prediction accuracy.

## Supporting information

Supplemental Methods and Results

## Data Availability

all data produced are available online at

https://dash.nichd.nih.gov/study/424597

