## Supplemental Methods and Results for "Predicting Walking Capacity Outcomes After Moderate to High Intensity Locomotor Training in Chronic Stroke"

### SUPPLEMENTARY METHODS

#### PRIMARY PROGNOSTIC STATISTICAL ANALYSIS

All prognostic analyses were done with an intent-to-treat approach using a linear model for 6MWD with fixed effects for group (HIIT or MAT), categorical testing time point (PRE, POST-4WK, POST-8WK, POST-12WK) and group-by-time interaction, with unconstrained covariance between repeated measures within participants.

6MWD ~ Group + Time + [Group x Time] + baseline FM-LL+ [baseline FM-LL x Time] + baseline CGS + [baseline CGS x Time] + baseline ABC + [baseline ABC x Time]

The prognostic value of different baseline covariates was assessed by adding fixed effects for the covariate(s) and their interaction(s) with time to the model above. The contrasts of interest were the baseline covariate-by-time interactions for each POST time point minus PRE. To obtain a standardized measure of prognostic model accuracy, a pseudo-$R^2$ statistic was obtained for each model and change time point. Pseudo-$R^2$ was calculated as the squared correlation between observed and model-estimated 6MWD changes at a given POST time point. Although the models estimated 6MWD changes for all participants (even those with missing data at the time point of interest), pseudo-$R^2$ could only be calculated using among participants with observed (i.e. Non-missing) values at a given time point. To obtain unstandardized measures of prognostic model inaccuracy in 6MWD meters, means and standard deviations of absolute prediction errors were also calculated based on the absolute difference between the model-estimated and observed 6MWD changes available at each time point.

Two highly implausible 6MWD values at POST-4WK were determined to be likely data entry errors and were set as missing. These 6MWD values were each >100 meters lower than the corresponding PRE value and >200 meters lower than subsequent POST values for those participants. 6MWD values at PRE, POST-4WK, POST-8WK and POST-12WK for participant 37 were 400, *272*, 558 and 549. For participant 55 they were 383, *250*, 468 and NA (missing). The implausible POST-4WK values underlined above were included in the previous analysis of this study to maximally control risk of bias for the between-group outcome comparisons, since both participants were in the HIIT group. These two values were removed from the current analysis because the risk of bias from doing so was determined to be lower for our current estimates of interest.

##### Preliminary analysis to assess between-group differences in prognostic effects

The overall modelling strategy described above assumed that the prognostic effects of a given baseline covariate were the same for both training intensity groups (i.e. No covariate-by-group-by-time interaction) on the population level. In other words, the main analysis strategy tested for general prognostic factors rather than predictors of differential treatment response to HIIT vs. MAT. Differential response prediction did not seem likely given the similarity between the intervention groups (aside from intensity dose), and we did not expect to be well powered for such an analysis based on the relatively modest sample size. Nonetheless, we still attempted it to assess the plausibility that the covariate effects were the same between groups.

This analysis added covariate-by-group-by-time interactions to the primary model and estimated the covariate-by-time interactions for each group for comparison. To help interpret the magnitude of the estimated effects, the coefficients were also scaled to a minimal clinically important difference (MCID) for the baseline covariate, to estimate the expected difference in 6MWD changes for a "meaningful" difference in the covariate within each group. This was done by multiplying the coefficients and their confidence intervals by the MCID. Then, these values were compared between groups, with reference to published MCID values for 6MWD.

### Exploratory prognostic analysis

This exploratory analysis tested all possible combinations of the potential baseline covariates (i.e. the all-possible regressions procedure[1]), with up to 5 covariates and their interactions with time in a given model. To allow fair comparisons between models, two participants with missing values for some baseline covariates were omitted from pseudo-$R^2$ calculations (but not model estimation) for all exploratory models. The prognostic value of each potential baseline covariate was estimated by the pseudo-$R^2$ contribution ($\Delta$ pseudo-$R^2$) from that covariate (and its interaction with time) to the model, pooled across all the models that included a given covariate. Each covariate was included in the same number of models. The $\Delta$ pseudo-$R^2$ of a given covariate in a model was calculated as the pseudo-$R^2$ for that model minus the pseudo-$R^2$ for a reduced model with all the same terms except those involving that covariate. Any small negative values, that occurred because pseudo-$R^2$ could only be calculated using among participants with observed values, were set to zero.

#### Variable importance analysis

The variable importance analysis compared the prognostic value ($\Delta$ pseudo-$R^2$) of different baseline covariates *within the same construct* (e.g. Different measures of gait speed), *between constructs (main part of this analysis)*, and *between change time points (*POST-4WK, POST-8WK, POST-12WK) for each construct. This required three separate parts to the variable importance analysis.

For the first part of the analysis we compared the prognostic value of *different baseline covariates within the same construct* (e.g. Different measures of gait speed) to guide covariate selection for each construct. A linear model was obtained for $\Delta$ pseudo-$R^2$ for each construct (averaged across the three change time points) from all models including a given covariate within that construct. The model estimated the mean $\Delta$ pseudo-$R^2$ for each covariate while allowing variance in $\Delta$ pseudo-$R^2$ to differ between covariates. Tukey-corrected p-values were obtained to account for the number of pairwise comparisons between covariates within that construct, when there were more than two covariates in a given construct.

The second and main part of the variable importance analysis compared the prognostic value of *different baseline covariates between constructs*. This main part of the analysis only included a single covariate for each construct, to manage collinearity and have a fairer comparison between constructs. For example, the mean $\Delta$ pseudo-$R^2$ for covariates representing a given construct could be artificially decreased by including more variables for a different construct that shared explanatory variance with the given construct. Thus, this analysis only included data from models that *exclusively* involved the single covariates selected to represent each construct (and no additional variables for that construct).

To have some protection against overfitting, these covariates were selected *a priori* as much as possible, including total FM-LL score for the motor impairment construct, CGS for gait speed, pain-limited walking duration (yes/no) for the pain construct, and one of the walking-related variables for the recent exercise history construct.[2] Remaining decisions about which covariate to use for each construct were based on which covariate had the highest $\Delta$ pseudo-$R^2$ in the first variable importance analysis above. For each change time point (and for the average $\Delta$ pseudo-$R^2$ across time points), a linear model was obtained to estimate the mean $\Delta$ pseudo-$R^2$ from that time point for each covariate. Variance in $\Delta$ pseudo-$R^2$ was again allowed to differ between covariates, and Tukey-corrected p-values were obtained to account for the 28 pairwise comparisons between covariates.

The third part of the variable importance analysis compared prognostic value *between change time points* within each baseline covariate. This part of the analysis included the same variables and data as the second part above. For each covariate, a linear model was obtained to estimate the mean $\Delta$ pseudo-$R^2$ from each change time point for that covariate, with unconstrained covariance between time points, and Tukey-corrected p-values to account for the three pairwise comparisons between time points.

Model selection

Starting from the reduced model set in the second (main) part of the variable importance analysis, we selected the 'best' model as the one that maximized average pseudo-$R^2$ across time points. The models with the highest $\Delta$ pseudo-$R^2$ were identified among those with the same number of covariates. The 'best' overall model was then selected as the one with the highest adjusted $\Delta$ pseudo-$R^2$, which was adjusted for the number of covariates in the model, using the formula: 1 - ( (1-pseudo-$R^2$) * N / (N – k – 1) ), where N was the number of participants contributing to the pseudo-$R^2$ measure, and k was the number of covariates in the model. Single covariate models were also obtained for each covariate to facilitate interpretation of the multivariable models.

### SUPPLEMENTARY RESULTS

#### PRIMARY PRE-SPECIFIED ANALYSIS

Preliminary analysis to assess between-group differences in prognostic effects

When temporarily expanding the primary model above to allow the prognostic covariate associations to vary by treatment group (i.e. Group-by-covariate-by-time interactions), the estimated 8-week 6MWD changes associated with an MCID difference in baseline FM-LL (6 points) for the HIIT and MAT groups were 31.3 and 23.6 meters respectively; a 7.8-meter difference [95% CI: -37.3, 52.8]. For an MCID difference in baseline CGS (0.10 m/s), these estimates were -3.2 and -4.6 meters; a 1.3-meter difference [-11, 13.7]. For an MCID difference in baseline ABC score (15%), these estimates were 11.6 and 10.6 meters; a 1.0-meter difference [-20.5, 22.5]. Since these between-group differences were all well below the 20-meter MCID for 6MWD (and not statistically significant), no further analyses attempted to estimate prognostic effects separately by group.

#### EXPLORATORY ANALYSIS

The 'all-possible regressions' procedure tested the 584,934 possible models with up to 5 baseline covariates (and their interactions with time), and 584,233 (99.9%) of these models converged. Each potential covariate was included in 74,482 of these models. The number of converged models that included a given covariate ranged from 73,886 (99.2%) to 74,482 (100%). Covariates sharing the same number of non-converged models were all in the same construct, indicating that non-convergence was likely due to collinearity from having highly correlated covariates in the same model.

Variable importance analysis

The first part of the variable importance analysis compared prognostic value (mean $\Delta$ pseudo-$R^2$ across change time points) between different baseline covariates within the same construct (supplementary figure S1, supplementary table S2). In the motor impairment construct, total FM-LL score (the *a priori* covariate) had significantly higher $\Delta$ pseudo-$R^2$ than FM-LL synergy score ($p<.0001$). In the gait speed construct, CGS (the *a priori* covariate) had significantly higher $\Delta$ pseudo-$R^2$ than most other variables in this construct ($p<.0001$), except CGS dichotomized ≥ 0.4 m/s and FGS, which both had significantly higher values ($p<.0001$). In the walking independence construct, FAC had significantly higher $\Delta$ pseudo-$R^2$ when dichotomized at > 2 vs. > 3 ($p<.0001$). In the assistive device construct, any assistive device had significantly higher $\Delta$ pseudo-$R^2$ than a weight bearing assistive device ($p<.0001$). In the pain construct, pain-limited walking duration (the *a priori* covariate) had significantly higher $\Delta$ pseudo-$R^2$ than pain severity or pain increases with walking ($p<.0001$). In the recent exercise history construct (where walking-related exercise history was the *a priori* category), walking exercise > 2 d/wk had significantly higher $\Delta$ pseudo-$R^2$ than most other covariates ($p<.0001$), except seated exercise > 0 d/wk, which had significantly higher values ($p<.0001$).

The second and main part of the variable importance analysis only included a single baseline covariate in each construct and compared prognostic value between different covariates/constructs (Figure 2). This analysis used data from the 12,615 possible models that *exclusively* involved the covariates selected to represent each construct (and no additional variables for that construct). All these models converged, and each covariate was included in 3,214 models. Most covariates had significantly ($p<.05$) higher $\Delta$ pseudo-$R^2$ than all covariates below it, except as indicated with matching superscript numbers in supplementary table S3.

The third part of the variable importance analysis compared prognostic value between change time points within each baseline covariate (supplemental figure S2), using the same data as the previous analysis. Most covariates had significantly ($p<.05$) different $\Delta$ pseudo-$R^2$ values between all-time points, except as indicated with matching superscript letters in supplementary table S3.

### SUPPLEMENTARY FIGURES

**Supplemental Figure S1. Prognostic value comparisons between baseline covariates within the same construct.** Left: Boxplots show the distribution of $\Delta$ pseudo-$R^2$ for each covariate across models to depict which covariate(s) in each construct explain the greatest variance in 6-minute walk distance change (Δ6MWD). Right: Boxplots show the distribution of T-statistics, primarily to depict whether each covariate was associated with greater (positive T-statistic) or lesser (negative T-statistic) Δ6MWD. Both: Values were averaged across time points within each model. Abbreviations: FM-LL, Fugl-Meyer lower limb motor scores; FAC, Functional Ambulation Category; ex, exercise; d/wk, days per week.

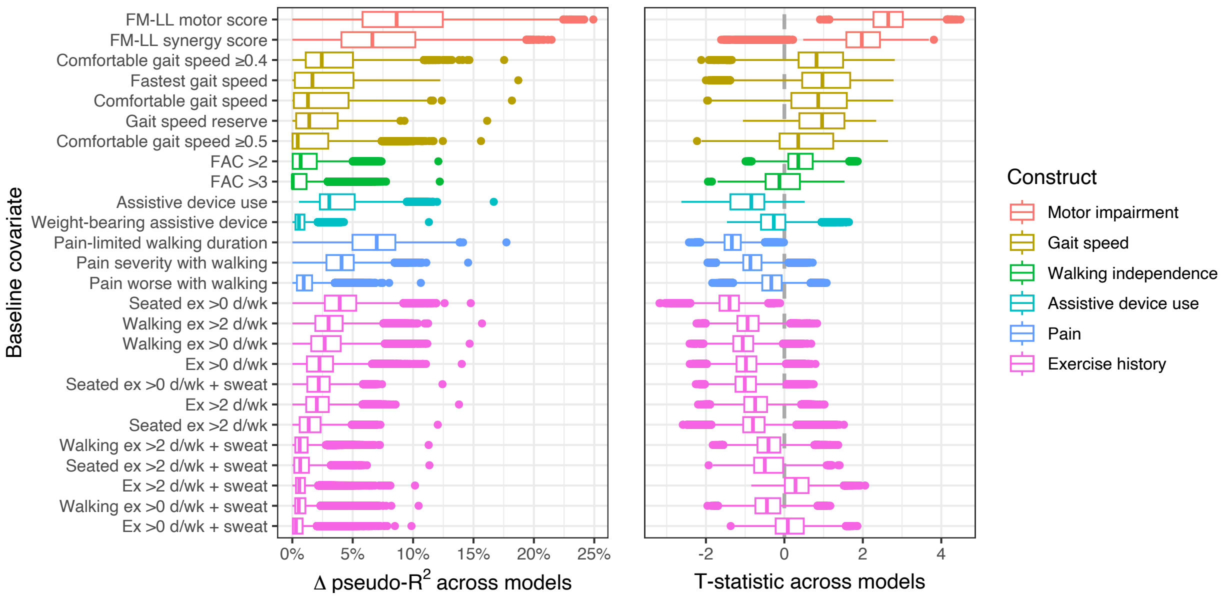

**Supplemental figure S2. Prognostic value comparisons between different change time points.** Top: Boxplots show the distribution of Δ pseudo-$R^2$ at each change time point across the 3,214 models involving each baseline covariate, to depict at which time point(s) that covariate explained the greatest variance in 6-minute walk distance change. Bottom: Boxplots show the distribution of T-statistics, primarily to depict whether each covariate was associated with greater (positive T-statistic) or lesser (negative T-statistic) changes in 6-minute walk distance at each time point. Abbreviations: FM-LL, Fugl-Meyer lower limb motor scores; ABC, Activities Balance Confidence Scale; PHQ, Patient Health Questionnaire; ex, exercise; d/wk, days per week; FAC, Functional Ambulation Category; mo, months.

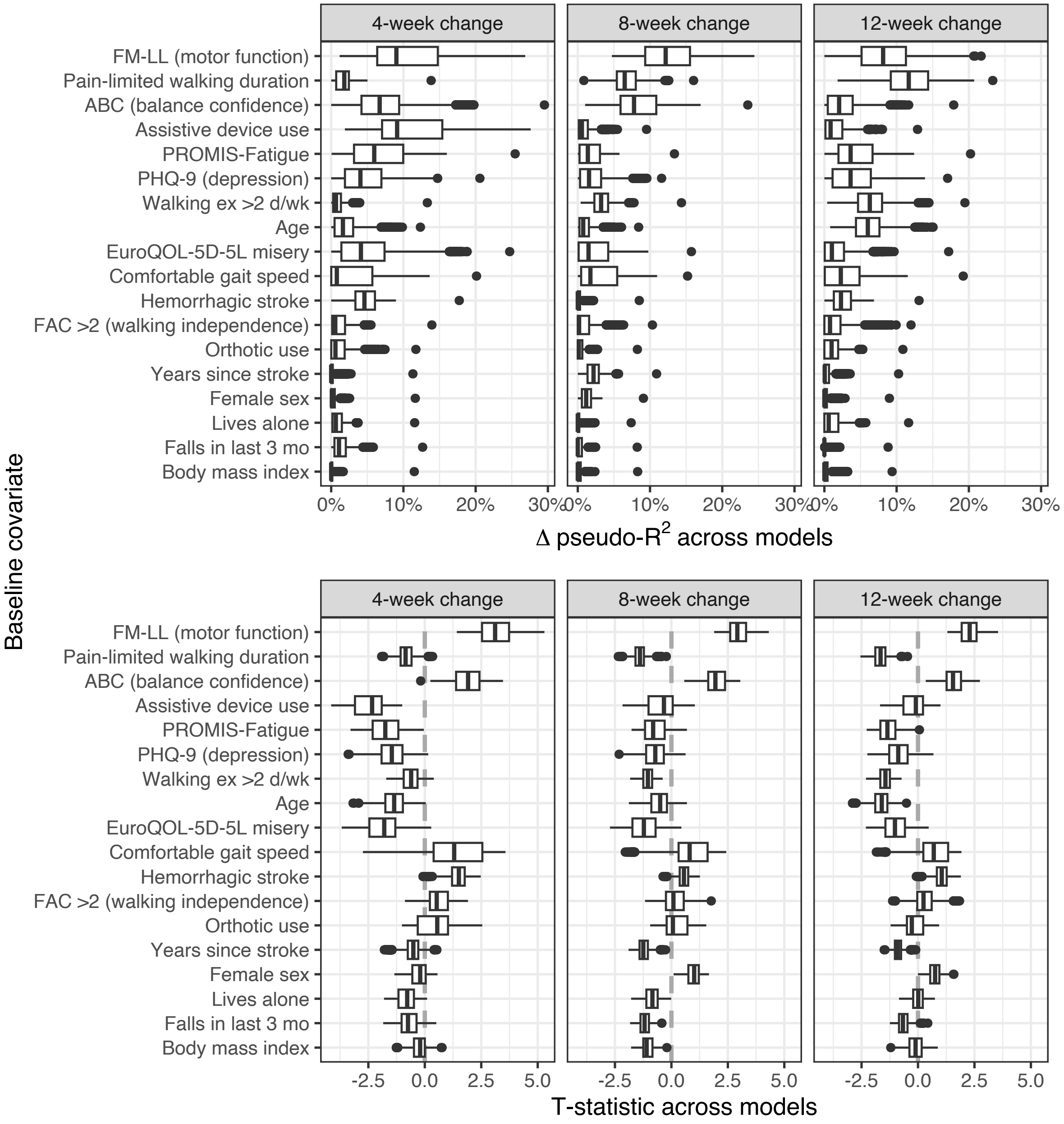
